# Identifying Challenges and Enablers to Engaging Patients in Preclinical Laboratory Research

**DOI:** 10.1101/2025.09.19.25336090

**Authors:** Madison Foster, Dean A. Fergusson, Emily Thompson, Victoria Hunniford, Talston Scott, Stephen Daniels, Dawn P. Richards, Pat Messner, Kathryn Hendrick, Patrick Sullivan, Asher A. Mendelson, Kimberly F. Macala, Kirsten M. Fiest, Angela M. Crawley, Bernard Thebaud, Stuart G. Nicholls, Cheryle A. Séguin, Grace Fox, Justin Presseau, Manoj M. Lalu

## Abstract

**Background:** Patient engagement in research enriches study design, conduct, and dissemination by integrating lived experiences of patients into the research process. Although patient engagement is increasingly common in clinical research settings, it remains rare in preclinical (i.e. laboratory based) research. To explore this gap, we conducted an interview study to understand how early adopters have implemented patient engagement in this area, focusing on the challenges and benefits of their approach.

**Methods:** We conducted semi-structured interviews of patients (n=15) and researchers (n=14) with previous preclinical patient engagement experience. Interviews were transcribed and conducted using an inductive, thematic content analysis, which allowed for bottom-up analysis of interview data. Our team inclusive of patients, clinical, preclinical and patient engagement researchers identified, reviewed, and refined emerging themes.

**Results:** We identified five themes. Researchers and patients highlighted the necessity to *adopt a thoughtful and tailored approach* for each preclinical engagement initiative (Theme 1). Clear communication was deemed critical, suggesting the need for a *clear and shared vocabulary* without technical jargon (Theme 2). This includes taking time to cultivate personal relationships, co-develop engagement activities to meet patient and researcher preferences and needs. In addition, v*aried goals for engagement in preclinical research* between researchers and patients was underscored (Theme 3), indicating the need to discuss aims and motivations early and often as well as to co-develop mutually beneficial strategies. Researchers and patients also discussed how they require *a better understanding of the value of preclinical patient engagement* (Theme 4). This could be fostered through education and illustrative case examples. Finally, a *shift in research culture* was deemed necessary (Theme 5), and called for stronger institutional support, efficient channels to connect preclinical researchers and patients, as well as initiatives that recognize and champion preclinical patient engagement.

**Conclusion:** Our study identified five common themes in preclinical patient engagement which can help the research community facilitate meaningful engagement of patients in preclinical laboratory research.

**Funding:** This work was supported by a Canadian Stem Cell Network Ethical, Legal and Social Implications Operating Grant. Patient engagement was supported by a Canadian Institutes of Health Research (CIHR) Strategy for Patient Oriented Research Catalyst Grant: Patient-Oriented Research. MML is supported by The Ottawa Hospital Anesthesia Alternate Funds Association, a University of Ottawa Junior Research Chair in Innovative Translational Research as well as the Canadian Anesthesia Research Foundation funded Canadian Anesthesiologists’ Society Career Scientist Award. AAM is supported by the Manitoba Medical Services Foundation Dr. F. W. DuVal and John Henson Clinical Research Professorship.

**Plain English Summary:** Engaging patients as partners in clinical research, known as patient engagement, is a growing practice that has numerous benefits. However, uptake in preclinical laboratory research (e.g. cell and animal studies) has been limited. Nevertheless, incorporating patients as active collaborators at this discovery stage of biomedical research may be beneficial. To better understand how patient engagement fits into preclinical research, we conducted interviews with patient partners and preclinical researchers who have been early adopters of this practice. Five key themes emerged. First, both groups emphasized the need for adopting a thoughtful and tailored approach since preclinical research is not typically patient facing. Second, shared vocabulary was important to facilitate communication. Third, setting clear expectations and outlining varied goals for engagement was considered critical. Fourth, understanding the value of preclinical research helped ground engagement efforts. Finally, interviewees felt a cultural shift is needed for this practice to be accepted more widely. These themes are important factors to consider when engaging patients in preclinical laboratory research; they may be used to inform and support future preclinical patient engagement efforts.

## Introduction

Patient engagement in research is the practice of integrating people with lived experience including patients and caregivers as collaborators or partners in the research process (see Box 1 for definitions).^1^ This practice has gained considerable traction^2^ and funding support^3–5^ in clinical research, in part due to increased recognition that patients should be involved as they are the “ultimate end-users” of biomedical research knowledge.^6, 7^ There is growing evidence demonstrating the many benefits to engaging patients as partners in clinical research, including ensuring appropriateness of procedures and patient-facing materials, enhanced relevance, improved study recruitment and dissemination, empowerment of patients, and increased public acceptance of novel scientific procedures.^8,6, 7, 9–12^<colcnt=1>

### Box 1.

**Patient Engagement Terminology**

#### Patient Engagement

*“Meaningful and active collaboration between researchers and patients in governance, priority setting, conducting research and knowledge translation. Depending on the context, patient-oriented research may also engage people who bring the collective voice of specific, affected communities.”*^13^

*“Research carried out ’with’ or ’by’ members of the public rather than ’to,’ ’about’ or ’for’ them"*^1^

Synonymous International Terms: Patient and Public Involvement, Consumer Involvement, Patient and Public Involvement and Engagement.

#### Patient

*“An overarching term inclusive of individuals with personal experience of a health issue and*

*informal caregivers, including family and friends.”*^13^

#### Preclinical patient engagement

*“Refers to engagement of patients in laboratory research, often*

*conducted using non-human animals, cells.”*

Engaging patients as partners in preclinical laboratory-based research, which is often conducted using animals or cells may hold similar benefits to those of clinical research, however published examples are rare and best practices for ‘preclinical patient engagement’ have not been formally established.^14^ Similar to clinical research, patients will be the ultimate end-users of preclinical research discoveries; thus, their perspectives can provide important insights that enhance research relevance. For example, patients have highlighted discrepancies between animal models used in research (i.e. and) and humans who live with the health conditions studied condition of interest (e.g. males and females, older people with co-morbidities)^15^. Similarly, certain clinical manifestations cannot be well replicated in animal models, leading to gaps in patient-prioritized preclinical knowledge and evidence. Exposing lab-based researchers to the ‘lived-experiences’ of patients who live with the health condition they study, may help start a dialogue between patients and scientists, allowing for a better understanding of patient experiences, and promoting integration of patient priorities into preclinical research.

Despite the many potential benefits of preclinical patient engagement, uptake has been limited. This was highlighted by a scoping review conducted by our team that identified only 32 published examples of patient engagement in preclinical settings. While this review demonstrated the feasibility and benefits of this practice,^14^ we noted a number of challenges to preclinical patient engagement including barriers in scientific jargon and a lack of guidance. In the current study, we further explored how these early adopter patients and researchers have worked together to overcome these challenges. This involved in-depth interviews of researchers and patients, identified in part through our scoping review. We aimed to capture their experiences, attitudes and perspectives regarding the engagement process. As this is an evolving area of practice, we chose to use an inductive approach to identify emerging themes.^16^ Our findings will help identify emerging best practices and inform the development of tools and resources to promote preclinical patient engagement.

### Objective

To better understand challenges and benefits encountered or anticipated by early adopters of preclinical patient engagement.

## Methods

### Study Design

Our team conducted a semi-structured, qualitative interview study.^16^ Results are reported as per the Consolidated Criteria for Reporting Qualitative Research^17^ (COREQ, Additional File 2) and Guidance for Reporting Involvement of Patients and the Public Short Form^18^ (GRIPP2-SF, Additional File 3) reporting guidelines.

### Ethics Statement

This study was approved by the Ottawa Health Science Network Research Ethics Board (Protocol 20200355-01H).

### Interview Guide Development

We developed separate interview guides for researchers (Additional File 4) and patient partner participants (Additional File 5). To develop questions, we reviewed clinical patient engagement frameworks^19–24^ and a previous interview study of patient engagement in health research.^25^ We generated broad questions aiming to elicit potential challenges and benefits of preclinical patient engagement. Both interview guides were developed with input from team-member patient partners and pilot tested with patient partners and researchers.

### Recruitment

We initially recruited using a purposive sampling approach by inviting corresponding authors of articles identified in our scoping review^14^ by email. Up to two reminders were also emailed. We then asked authors who participated to refer additional team members or contacts who might be interested and eligible to participate. We used the 10+3 rule,^26^ whereby we conducted at least thirteen interviews of patients and 13 interviews of researchers to assess data adequacy. Data adequacy was deemed sufficient when no new major themes emerged in the last three consecutive interviews.

### Interview Procedure

All participants provided verbal informed consent. A trained and experienced clinical research assistant (MF) conducted individual interviews over videoconference calls that were recorded. A contracted transcriptionist transcribed the recordings verbatim. Personal identifying information was removed. We then imported transcripts into a qualitative software program NVivo 11 (Lumivero, Denver, Colorado) for analysis.

### Data Analysis

The steps for thematic content analysis^16^ informed our process for identification of key themes. This inductive analysis approach^16^ allows for the development of key themes from the interview data (also referred to as a ‘bottom up’ or ‘data-driven’ approach), rather than categorization by an existing framework or theory (a ‘top down’ approach). First, the research assistant who conducted all interviews developed an initial draft codebook through review and familiarization of the transcripts. Three research assistants (MF, VH, ET) then coded six interviews in duplicate. Consensus meetings were held between coding of every two transcripts to refine the codebook. Team feedback from the co-authors was also sought iteratively throughout this process. Once the codebook had been finalized, one research assistant (MF) coded the remaining researcher interviews, and one research assistant (ET) coded the remaining patient interviews. We generated themes iteratively through review of codes and text excerpts, discussion between coders, and discussion with all team members (i.e., patients and researchers). We then generated global themes (e.g. representative of both populations) through discussion between coders and using feedback from team members.

### Research Team and Reflexivity

Our team included early, mid and senior career stage researchers, some of whom are also clinicians, with expertise spanning preclinical, clinical and patient engagement research (MML, DAF, KFM, AAM, AM, KF, AC, BT, CS, DPR). The team also encompasses a knowledge translation scientist and health psychologist (JP), a patient-oriented research facilitator (SGN, DPR), patient partners (DPR, KH, PM, PS) and highly qualified personnel (research assistants; ET, VH, GF, MF, TS, SD). Team feedback from all co-authors was sought throughout the entirety of the research process.

Our primary interviewer and analyst (MF, female, M.Sc. Epidemiology) had prior experience conducting and analyzing qualitative interview studies to identify challenges and enablers.^27–31^ Due to purposive sampling in a field of study that our team has networked within (i.e. preclinical patient engagement), the interviewer had previously met some of the interviewees in a professional capacity prior to study commencement.

### Patient Partner Involvement

Our team includes four patient partners (DPR, PM, PS, KH), who were compensated in alignment with Canadian Institutes of Health Research best-practice guidelines. DPR and KH were involved throughout the research project, including the development of grant applications that funded this work, applying to the research ethics board, preparing recruitment materials, and contributing to the interview guide. Patient input helped to ensure recruitment materials were approachable and clear. PM and PS aided in the piloting of the patient interview guide, to provide the research assistant with feedback on question framing, wording and flow. All patient partner team members attended team meetings to discuss interview study findings and provide feedback on identified themes. All patient partners have also contributed to this manuscript as co-authors.

## Results

### Participant Characteristics

In total, thirty interviews were conducted (16 patients, and 14 researchers). During coding, we excluded one patient interview from analysis as we determined they had engaged in clinical (not preclinical) project. **Table 1** describes researcher and patient participant characteristics. Our sample represented a diverse range of research areas, career stages, and life perspectives. The patient engagement initiatives that participants discussed are briefly summarized in Supplemental Table 1 (Additional File 1).

**Table 1.** a) Researcher Participant Characteristics.

| **Characteristic** | **Number of participants**<br>**or median (range)** |
| --- | --- |
| **Number of Participants** | 14 |
| **Gender** |  |
| Women | 7 |
| Man | 7 |
| **Area of Research (Health Condition/Disease)** |  |
| Musculoskeletal system/ connective tissue | 4 |
| Mental & behavioral disorders | 3 |
| Nervous system | 3 |
| Certain infectious & parasitic | 1 |
| Endocrine, nutritional & metabolic | 3 |
| Genitourinary system | 1 |
| Cancer | 2 |
| Genomics | 1 |
| **Research Career Level** |  |
| Early (0-5 years as independent researcher) | 3 |
| Mid (5-15 years) | 5 |
| Senior (15+ years) | 5 |
| Unspecified | 1 |
| **Median Age (range)** | 50 (34-54) |
| **Median Interview Length in Minutes (range)** | 62 (49-81) |

**Table 1.** b) Patient Participant Characteristics.

| **Characteristic** | **Number of participants**<br>**or median (range)** |
| --- | --- |
| **Number of Participants** | 15 |
| **Gender** |  |
| Woman | 8 |
| Man | 7 |
| **Perspective** |  |
| Lived experience of a health condition | 10 |
| Caregiver | 4 |
| Both | 1 |
| **Everyday Life (i.e. Roles and Relationships)** |  |
| Work full-time or part-time | 6 |
| Patient partner in research, patient-advocate | 5 |
| Caregiver (parent, sibling) | 5 |
| Retired | 3 |
| Student | 1 |
| **Median Age (range)** | 58 (25-79) |
| **Median years involved in research as a partner (range)** | 9 (1.5-21) |
| **Median Interview Length in Minutes (range)** | 69 (43-81) |

**Table 2:** Identified themes and key take-aways.

| Theme | Key Take-Aways | Illustrative Quotes |
| --- | --- | --- |
| 1. Need to take a thoughtful and tailored approach | Involve patients early and throughout the research project; establish a relationship and keep lines of communication open<br>Co-develop the engagement approach, taking patient partner needs, preferences, potential burden and contributions into account<br>Actively aim to avoid tokenism | *"[...] we do it in what I would hope is a thoughtful way, but not a very structured way." – R7*<br>*"There are no rules the only rule is if you want to know something of the patients, ask [...] of course, that goes for patients, if you want to have questions the first point to ask is really your researcher and go to the team–" – PP1* |
| 2. Clear and shared vocabulary | Work together at the beginning of the engagement initiative to define key scientific terms and co-develop resources (e.g. glossary)<br>Ensure effort is made to explain the research in non-technical language and encourage questions and comments | *"[...] there are two challenges [...] understanding the project that you're working on especially if it's really heavy, heavy science or high tech or something. Secondly, is actually understanding how research operates perhaps particularly in one of the therapeutic areas or whatever technological area [...]" – PP10*<br>*"And, and I guess that's one structure to, to probably think about early on and how you kind of how you can work towards a common language." – R3* |
| 3. Varied goals for engagement in preclinical research | Discuss aims for both patients and researchers to identify activities that are valuable and beneficial to both groups<br>Consider that partners may be especially interested in downstream implications of the research; this can help to provide new perspectives, and identify new and unique activities and areas for involvement | *"[...] we're not in research to be basic scientists. We're mainly there so that, you know, we see the safety and benefit of research for patients." – PP3*<br>*"[...] knowing what we were trying to get out of it really helped. Lots of people go with these and go oh I need to involve patients and do it without actually having a goal orientation in it." – R1* |
| 4. Understanding the value of patient engagement in preclinical research | Education and case studies may help to change individual mind-sets and clarify perceptions hindering patient engagement (e.g. patients not interested in basic science, no benefits) | *"[...] a lot of reasons that are widely reported [...] preclinical researchers struggle to see the benefits and the impact and the need to [involve] people in their, meaningfully in their research as compared to care researchers [...]" – R10*<br>*"[...] when a patient advocate first sits down at the table with especially a basic scientist (laugh) there's not an immediate oh I need you (laugh) [...] you more or less have to show how you can help them because they've got [...] a tube focus and I understand that." – PP3* |
| 5. Shift in scientific culture is required | Changes to research culture (e.g., provision of guidance, funding resources, training and endorsement by institutions) may help to encourage engagement of patients in preclinical research | *"[...] I want to again highlight the importance of having sort of a maybe general change of culture in research [...] it cannot happen by just a few again researchers taking the lead." – R5*<br>*"[...] in the academic world working with patients is not always promoted or given much credence." – PP2* |

From our analysis of interviews, we identified five themes 1) Need to Take a Thoughtful and Tailored Approach; 2) Clear and Shared Vocabulary; 3) Varied Engagement Goals in Preclinical Research; 4) Understanding the Value of Patient Engagement in Preclinical Research; 5) A Shift in Scientific Culture. Below we describe each in greater detail.

### Theme 1: Need to Take a Thoughtful and Tailored Approach

The first theme recognized the need to take a thoughtful and tailored approach to the engagement process. While our interviewees underscored that there is *no generalized formula for engaging patients in preclinical research, there is a need to co-develop and tailor the engagement to the project or initiative, while also allowing for flexibility*. This approach helps mitigate tokenistic inclusion of patients and unilateral decision making on behalf of the research team, allowing engagement activities and roles to develop organically and through discussion. Like guidance in clinical settings, the *importance of establishing a partnership and creating a safe, comfortable and trusting environment/team dynamic* and maintaining open dialogue were noted. As patient engagement in preclinical research is still new to many, some expressed that it is a learning process for both patients and researchers.

> *“The key point is that there’s no one size fits all […]. Because yes they may not be familiar or comfortable with reviewing grants, but they might be able to contribute in other aspects and that is absolutely fine. I think to give them [the] opportunity to see that [like] any other thing in research […] it’s a team [effort]. So a team requires every aspect to be addressed and if they feel comfortable to contribute to one or two areas that is fine […] I think it is important to remind them about the value of that.”* - Researcher 5

One key message expressed throughout the interviews was the need to *involve patients as early as possible in a research project.* This may help them to become familiar with a project and allow them to provide even more informed insights, as opposed to the frustrations expressed by patients about being brought into a study late in the processes.

> *“I was brought into the project while it was already underway with very little introduction. Sort of like sink or swim figure it out for yourself kind of thing … This was something that was very foreign to me, so the learning curve was extremely steep”* – Patient 14

> *“We saw a benefit [in] involving people across the whole of the research cycle … Having continuity and having the same people involved rather than getting different people involved at each stage … was valuable because sometimes preclinical research is harder for the volunteers to get their head around. It needs a bit of time and support for them to understand it. Having that length of involvement means there’s more value to it.”* – Researcher 10

Both groups emphasized the importance of considering and respecting patient partner needs, preferences, potential challenges, and contributions. Patients also highlighted the necessity for researchers to offer the same level of respect and attention to patients as they would to other research collaborators, avoiding assumptions about patient interest and involvement. Patients especially placed importance on flexibility in selecting projects tailored to individual bandwidths and interests.

> *“There might have been meetings where the researcher had decided ‘I didn’t think that you were interested in this’. Instead of asking ‘we have this meeting, are you interested to attend or not?”* - Patient 1

They expressed the need for researchers to be open to partner ideas, allowing for patient-led initiatives and keeping discussions open. Co-developing and tailoring programs to align with patient preferences may help researchers recognize that patients do not follow the same timelines as their research counterparts and will require more notice for deadlines.

Overall, researchers’ accounts of patient engagement experiences were positive and found to be meaningful and rewarding. However, some researchers did stress the *preparation, time, and effort required for patient engagement*.

> *“I think some of the challenges that are experienced are because people go in under prepared.”* – Researcher 14

In the few negative examples shared by researcher participants, issues tended to relate to time/funding limitations, and lack of other researchers’ effort to involve them, or their preconceived notions (e.g. not understanding the value of patient engagement). Though patients often shared positive experiences, they also suggested that negative experiences can happen. While they saw such encounters as opportunities for growth, they also stressed *the need to actively avoid tokenism.* Negative experiences expressed by patients included publishing scientific articles that included patient names without their input, disregard for patient availability and priorities, making decisions without patient input, and projects being dropped without further insight or follow-up. Ensuring a level-playing field whereby patients are active members of the team and treated as such was underlined as important; it was noted that to achieve this, modification of timelines may be required.

### Theme 2: Clear and Shared Vocabulary

Both patient and researcher participants noted that preclinical research often involves specialized language and concepts that is often harder to explain and understand than clinical research. Although researchers felt responsible for using accessible vocabulary, interestingly patients believed it was their duty to ask questions and learn, with the researchers providing necessary support.

> *“I think it’s just to keep the lines of communication open and ask a lot of questions. Don’t be afraid to ask questions.”* – Patient 5

> *“I think it’s initially just making sure that the lines of communication are open and that anything that’s sent out about it or presented on the day is reviewed by non-researchers and members of the public and patients. I think that’s really, really important. And then if you’re obviously on a call and jargon comes up to feel confident to say I don’t know what that means I don’t understand it.”* – Patient 5

Both groups shared advice for working towards a shared vocabulary. One approach was to co-develop resources, consulting patients to ensure that language met their needs. Other practices described included information sessions, visual demonstrations, glossary of terms, focusing on broad concepts, using analogies, adopting an appropriate pace, describing projects or concepts in “smaller units”, and creating an environment conducive to asking questions.

> *“Sometimes there was a mismatch between what the researchers felt was clear and lay and what the volunteers felt is clear and lay. So often that would require us maybe to encourage the researchers to try and to use more accessible language sometimes or to go a little bit slower in their presentations in particular.”* – Researcher 10

### Theme 3: Varied Engagement Goals in Preclinical Research

This theme highlighted differences between patient and researcher motivations for patient engagement. Patients were *informed by a clinical past, looking towards an improved clinical future*. Many cited personal experiences as the catalyst for their involvement and were looking to see how their engagement could impact future patient care and research translation.

> *“I think patients are committed to nurturing the next generation of [health condition] researchers because we have a vested interest […] in better [health] care”* – Patient 2

In contrast, researchers reported varied goals for patient engagement. Some aimed to share knowledge - for example, helping pediatric patients understand preclinical research - while others prioritized exchanging perspectives to inform study design. Over time, many researchers shifted toward this latter goal, with programs that featured multiple levels of engagement, and a recognition that patient engagement could involve the entire preclinical research continuum.

> *“You have to [ask], what does the patient want out of this experience and what [do] I want out of it and how to bridge that gap […] And I don’t think it’s you’ll always stick to the same objectives - it’s something that will evolve with time.”* – Researcher 12

### Theme 4: Understanding the Value of Patient Engagement in Preclinical Research

The final two themes centered on evolving mindsets: how the participants came to value preclinical patient engagement, and howe broader shifts in scientific culture are needed to support and sustain this work. The first highlights the importance of understanding and emphasizing the value of engaging patients in preclinical research. This was deemed an integral component of how to promote meaningful preclinical patient engagement as it encouraged ‘buy-in’ from both researchers and patients, reducing the likelihood of tokenistic (i.e. a false appearance of inclusiveness”^2^) ‘check-box’ engagement practices. As identified through researcher interviews and highlighted by patients, preclinical researchers were highly motivated to engage patients, and patients were motivated to engage in the preclinical research process. Similar to patient engagement in clinical research, interviewees identified several benefits of preclinical patient engagement, including: a) improving research relevance by incorporating lived experience into study design and interpretation; b) humanizing preclinical research by connecting it to patient stories; c) helping patients learn about ongoing research; and d) helping researchers better understand the lived experience of the condition, which some found motivating for pursuing and prioritizing their research..

> *“[Patient engagement] gives you more of a meaning to your research in terms of a practical sense [of the eventual] impact it could have.”* – Researcher 12

However, some researchers noted that the benefits of patient engagement in preclinical research are less intuitive than those in clinical research. As a result, they suggested that other scientists have not recognized its value, leading to limited investment and suboptimal engagement efforts. For example, one researcher noted that (prior to their successful engagement work) they initially thought it was not their role to engage patients, and that community partners would not be interested in their preclinical work. Another researcher expressed initial concern that patient engagement may lead to inflated hope and expectations for a cure. Both later revised their views after working with patients. These examples highlight the need to address misconceptions through education and greater awareness of preclinical patient engagement.

> *“In terms of preclinical research, people often are studying [the] mechanisms and biological or scientific processes and not […often…] thinking about the big picture … Interacting with patients and hearing their experiences makes you much more aware of [the] issues and [how] you can factor that into your research approach.”* – Researcher 6

From the patient perspective, the benefits of being involved in preclinical research included a sense of empowerment and feelings that they were more assertive and knowledgeable about their health condition.

*“This work [is] both satisfying but also intellectually very challenging and interesting […] I know a lot more about [condition] by being involved with this and I really enjoy the education side […] We’re committed to this generational change and working with the students and post docs [to make] them comfortable in this world of working with patients.”* -Patient 2

Another highlighted benefit was bridging the gap between researchers and the public by making preclinical research more accessible. Several patients also alluded to the importance of contributing to improved care for future patients. Overall, both researchers and patients expressed enthusiasm for engagement and recognized the importance of highlighting the benefits of this practice.

> *“As a patient, I value the work … from start to finish. It’s the work that keeps patients like myself alive. So I find it extremely valuable and I love to contribute whenever I can.”* – Partner 9

### Theme 5: A Shift in Scientific Culture is Required

A second theme identified across interviews was the need for a shift in scientific culture. Both patients and researchers noted the novelty of engaging patients in preclinical laboratory research, and the importance of specifying how patients can be involved in the process. It was noted that this will require *training initiatives and resources* for both researchers and patients.

> *“I would have a hard time convincing them that what we did is valuable … We need to change the culture and bring this to attention at all levels otherwise we won’t be able to continue working on it.”* – Researcher 5

Additionally, both patients and researchers identified limited funding as a potential barrier. Researchers noted that funding was often lacking during the early, pre-grant stages when engagement could help shape research ideas. Other researchers noted that while they received funding specifically for patient or community engagement, they were limited by small budgets and no funding mechanisms to partner beyond a grant period. Patients also acknowledged challenges in securing funding.

> *“You have to start it in the preclinical. You have to get the best ideas there because the funding is very limited no matter what area you’re in.”* – Patient 3

These limitations in training and funding allocation highlighted the need for institutional and organizational involvement in the process to *create and maximize opportunities* that would shift scientific culture towards preclinical patient engagement. For instance, the initiation of partnerships between preclinical research organizations and community organizations, charities, research networks, and hospital networks were cited as enablers that could facilitate connections between patients and preclinical researchers. In addition, interviewees suggested there should be academic and/or funding rewards for preclinical patient engagement. Indeed, researcher interviewees suggested that the time required for preclinical patient engagement was neither recognized nor rewarded by institutes/funders.

> *“An academics career is really tough as it is, right? […] If you really want to encourage good practices, they should be rewarded.”* – Researcher 2

> *“The other thing that will help is that currently there’s not any career benefits to it, you know. […] I’m judged on short-term metrics and public involvement is not one of them.”* – Researcher 1

The need for *motivation at the organizational level* could help reconcile this by promoting educational, training and award initiatives that recognize and encourage preclinical patient engagement. Posited solutions included patient involvement in the evaluation processes (e.g. interview committees, advisory panels, grant review), requiring patient engagement in research proposal (for example, via the funding agency or ethics committee), as well as buy-in and encouragement from leadership/institutions.

It was recognized that one way of initiating cultural change was to target early career researchers and trainees by making it an integral part of their training.

> *“We’re committed to this generational change and working with the students and post docs [to make] them comfortable in this world of working with patients.”* – Patient 2

> *“[T]he only thing I think that we need to do is, is keep reaching out to the researchers especially the younger ones to let them know what the advantages are to have us onboard. […] having them understand that what you’re doing with mice you can’t always do to humans […] ”* – Patient 4

## Discussion

Through interviews with early adopters of preclinical patient engagement, we identified key challenges and benefits experienced by these early adopters, along with some potential factors that could support meaningful engagement. Overall, both patients and researchers shared positive experiences and motivations from their involvement, spanning a wide range of research topics. Given how little is published about preclinical patient engagement initiatives,^32^ it is clear that the researchers and patient participants in our study are ‘early adopters’. Practical considerations identified by our study include developing a shared vocabulary, tailoring engagement approaches, and working together to identify mutual goals. To cultivate the next wave of preclinical patient engagement, our findings suggest that increasing awareness about the benefits of patient engagement for both the researchers and patients, as well as encouraging institutional support for meaningful engagement initiatives backed by funding and career advancement opportunities

From a practical standpoint, some participants noted the challenge of knowing where to start with patient engagement initiatives. A promising practice identified by our interviews was for teams to take a *tailored approach*. Tailoring begins with establishing relationships, which has also been emphasized by others suggesting that ‘training is not a substitute for relationship building’ in preclinical patient engagement.^33^ Within health research, the importance of “co-building social relations” and “co-creat[ing] environments that minimiz[e…] risks” has also been recognized.^25^ This relationship building provides a foundation to tailor a project-specific engagement plan reflective of both researcher and patient partner interests, needs, and availability. Further, it facilitates open, transparent communication and the development of a shared vocabulary. In co-creating and tailoring engagement plans, teams should consider and incorporate guidance that is beginning to emerge around preclinical patient engagement.^34–39^

We also identified varied goals, aims and motivations for engagement in preclinical research, emphasizing the need to consider project-specific aims and person-specific perspectives. Initiatives typically fell into three broad categories: 1) educating patients about ongoing research, 2) building partnerships for mutual learning and shaping the research, or 3) co-designing the engagement initiative and supporting other patients partnering with the team. Participants noted that some initiatives started as one-way conversations and evolved into two-way dialogues. This shift reflects the move away from a ‘Deficit’ model, which assumes patients lack knowledge and need education, to a ‘Dialogue’ approach that values lived experiences and fosters two-way engagement.^40^ We propose that this evolution could be accelerated by early team discussions that address everyone’s aims, goals, and motivations for partnering, allowing for the identification of shared goals and activities.^41^

As noted, a broader cultural shift in preclinical research (i.e., a supportive climate for patient engagement) will be required for more widespread adoption. Although the need for similar shifts has been suggested in clinical research,^42^ we hypothesize that challenges due to knowledge gaps may be more pronounced in preclinical research. Patient engagement may be more intuitive in clinical research due to clinical researchers’ familiarity with providing patients with clinical care, experiences communicating with patients, and ample guidance on patient engagement. Difficulties incorporating patient engagement into preclinical research were also expressed in studies exploring challenges implementing patient engagement in lab-based spinal cord injury (SCI) research, with patients describing the gulf between the daily lived experience with SCI, and the lab-based research environment.^43^ Overcoming this gap helped patients become more informed and better equipped to contribute meaningfully, allowing their input to shape research priorities and outcomes early in the translational pipeline.^43, 44^ Our results suggest that resources and recognition could facilitate this cultural shift in preclinical research. Organizations could ensure appropriate resources are available for researchers and patients to collaborate (e.g. education, training modules,^45–47^ ^18^ and reporting guidelines). Moreover, incorporating patient engagement into calls for a ‘team science’ approach and rewarding the practice may also help raise awareness and motivation for this practice in preclinical laboratory research. Furthermore, as more funders and institutions adopt patient engagement in preclinical research, and as researchers are increasingly rewarded for this work, a broader cultural shift is likely: one where engaging patients becomes both a meaningful practice that impacts research and a recognized avenue for career advancement.

## Strengths & Limitations

Our semi-structured interview approach has been used in other preclinical patient engagement studies that investigated challenges and benefits of integrating patient engagement into SCI lab-based research.^43, 44^ While these studies focused on one specific disease state, our approach casts a wider net across a diverse range of health conditions; this complements previous research, and generalizes the findings to many different contexts. Our eligibility criteria required prior experience with preclinical patient engagement, which may have biased findings, rather than capturing the full range of barriers faced by those without such experience. This includes those who were unsuccessful in their initiatives, did not respond to our recruitment efforts, or have not yet attempted engagement. In addition, data interpretation and analysis may be influenced by team traits, characteristics, and inherent biases (e.g., believing patient engagement has intrinsic value). To address this, we have transparently reported on team characteristics as suggested by the COREQ reporting checklist.^17^ We followed suggested best practice for determining adequate sample sizes, and we believe our study provides rich accounts of both patient and researcher experiences in preclinical patient engagement.^26^

## Conclusion

Our interview study explores the perspectives of patients and researchers who have prior experience in preclinical patient engagement initiatives. In addition to identifying some potential challenges to patient engagement in this space, we have also identified several practical considerations for planning and implementing patient engagement in preclinical laboratory research. These insights will inform the development of educational guidance and resources to promote best practices and facilitate more widespread implementation of this practice.

## Supporting information

Supplementary Files - Identifying Challenges and Enablers to Engaging Patients in Preclinical Laboratory Research.

## Data Availability

Data are available in the main text or the Supplemental Material are available from the corresponding author on reasonable request.

## Contributors

Conceptualization: DAF, JP, MML, DPR, KH, AAM, KFM, KMF

Data curation: MF, ET

Formal Analysis: MF, ET, VH

Funding acquisition: DAF, JP, MML, DPR, KH, AAM, KFM, KMF

Investigation: MF

Methodology: DAF, JP, MML, DPR, KH, PM, AAM, KFM, KMF, MF, GF

Project administration: MML, MF Resources: DAF, JP, MML, DPR

Supervision: DAF, JP, MML Visualization: MF, ET

Writing – original draft: MF, MML

Writing – review & editing: DAF, JP, MML, DPR, KH, PM, PS, AAM, KFM, KMF, ET, VH, GF, MF, TS, SD, AMC, BT, SGN, CAS,

## Acknowledgements

This work was supported by a Canadian Institutes of Health Research (CIHR) Strategy for Patient Oriented Research Catalyst Grant: Patient-Oriented Research, as well as a Canadian Stem Cell Network Ethical, Legal and Social Implications Operating Grant. MML is supported by The Ottawa Hospital Anesthesia Alternate Funds Association, a University of Ottawa Junior Research Chair in Innovative Translational Research as well as the Canadian Anesthesia Research Foundation funded Canadian Anesthesiologists’ Society Career Scientist Award. AAM is supported by the Manitoba Medical Services Foundation Dr. F. W. DuVal and John Henson Clinical Research Professorship.

The authors would like to thank the interviewees for participating in their interview study.

## Data Sharing Statement

Data are available in the main text or the Supplemental Material are available from the corresponding author on reasonable request. Data and materials individual participants will not be shared to protect privacy.

## Supplementary Files

**Additional File 1:** Supplemental Table 1: Summary of Patient Engagement Initiatives by Research Phase

**Additional File 2:** COnsolidated criteria for REporting Qualitative research (COREQ) Checklist

**Additional File 3:** GRIPP2-SF Checklist

**Additional File 4:** Interview Guide

**Additional File 5:** Code Book

