## Supplementary Files - Identifying Challenges and Enablers to Engaging Patients in Preclinical Laboratory Research. for "Identifying Challenges and Enablers to Engaging Patients in Preclinical Laboratory Research": Additional file 1_Supplementary table.docx

**Supplemental Table 1: Summary of Patient Engagement Initiatives by Research Phase**

| **Phase** | **Initiative/Activity** | | |
| --- | --- | --- | --- |
| Priority Setting | - Partnered for priority setting (R5) | - Public provided comments online on guidelines developed for research. Researchers then incorporated comments into the guidelines. (R8) | - Took part in a larger initiative where patients organized a meeting for priority setting (R11) |
| Funding | - Involved in grant application for basic science, reviewing the patient engagement strategy (P1) - Grant review (P3, P10) - Grant development and writing (P3) | - Involved throughout the whole project, including funding (P4) - Research funder helped facilitate partnerships for both preclinical and clinical researchers (patient research monitors assigned to grant review). (R10) | - Partnered with families to fund research (R11) - Wrote letters of support for research teams (P9) |
| Education | - Built community of partners to work with on an ongoing basis. Worked with partners to develop an educational event for pediatric patients to learn more about basic science (R1). - Funder assigned patient ‘research monitors.’ Asked research monitors to participate in a video discussion with researchers on animal use. Disseminated to share conversation (R2). - Educational events – children/parents attended one time and had discussions with genetic researchers. Also completed surveys on attitudes towards genetic research (R4). | - Seminars initiated by patient organization. Now a formal course where patients attended lectures by students to learn about cancer research. Also, held team building activities to learn about the patient perspective. (R6) - Shared research with patient organizations, asked patients questions. Focus groups with educational session and discussion to learn patient perspectives on research. PhD student also engaged with a patient partner throughout their project. (R14) - Attended a scientific conference that had a program for patients to be partnered with a student (P11, P12) | - Wrote a grant and helped to carry out an initiative to raise awareness of tissue banking among patients (alongside researchers) (P3) - Worked with a researcher on a project to involve/raise awareness of research in young people (P5) - Described project to other patient groups/ helped to develop accessible materials (P10, P14) - Helped to develop accessible slides to describe the lab project (to patient research partners and for a public-facing website). (P15) - Lab tours (P10, P14) |
| Study Design | - Involved throughout whole project, including question, design, and getting additional collaborators involved. (P4) | - Involved in developing the consent forms and questionnaires to collect tissue samples for research purposes. (P15) |  |
| Exchange of Information/Perspectives | - Partnered for hypothesis generation & feedback throughout the study (R5) - Partner with ‘research participants’ (those who donate samples for research). Aim to understand their priorities and share research. (R7) | - Engaged patient partner in lab-based projects (e.g. team meetings, developed questions with patients, sought patient perspectives). (R12, R15, P6, P10, P14) - Shared & discussed information learned from research with families (R11) - Provides their perspective as a patient representative. Invited researchers to speak at their organization’s event. (P9) | - Research funder helped facilitate partnerships for both preclinical and clinical researchers (patient research monitors assigned to projects). (R10) - Engaged caregiver partner in lab-based projects (e.g. get together to discuss ongoing research and updates from the patient perspective). (R13) |
| Publication Phase of Research – Dissemination/Awareness | - Helps with dissemination (e.g. presentation of posters and speaker at a scientific conference) (P4, P6, P11) | - Conference that invites patients to attend. Also had a workshop with teens – asked what they wanted to be involved in – chose communication of ongoing research (R3). | - Planning to write thank you letters to those who donated tissue (P10, P14) |
| Supporting Engagement & Other | - Engagement facilitator for a laboratory project; helped to train other patients. (P1) - Helps to train other patient research advocates and researchers and conducts outreach. Also sits on an advocacy planning committee. (P4) - Program coordinator helps to bring patients onboard to review projects, be an advisor on a project, or be member of the Patient and Family Advisory Committee. (P13) | - Works with a member at a university to bring together basic science students and patients for seminars (now a formal course) to allow students to meet patients, and discuss their research in lay language. (P2) - Caregiver and researcher with specialization in ethics (on ethics board). Facilitated consultation with patient/public community. Involved two in-person events, and a survey to obtain feedback. (P16) - Helps plan patient engagement (P7, P10) | - Helped to develop recommendations for ‘patient research and collaboration’ or researchers/ patient research partners (P1, P15) - Helped with an interview study to assess engagement for a laboratory project (P1) - Co-author (P1) - Lay judge for a multidisciplinary problem-solving competition workshop (e.g. focused on working towards treatment for a specific cancer) (P6) |
