## Supplementary Files - Identifying Challenges and Enablers to Engaging Patients in Preclinical Laboratory Research. for "Identifying Challenges and Enablers to Engaging Patients in Preclinical Laboratory Research": Additional file 2._ COREQ.docx.docx

**Supplemental File X: Supplementary COnsolidated criteria for REporting Qualitative studies (COREQ) checklist**

| **No. Item** | **Guide Questions/Descriptions** | **Reported on Page #** |
| --- | --- | --- |
| **Domain 1. Research team and reflexivity** | | |
| **Personal Characteristics** | | |
| Interviewer/facilitator | Which author/s conducted the interview or focus group? | MF, AC |
| Credentials | What were the researcher’s credentials? E.g. PhD, MD | MSc, BSc  1 |
| Occupation | What was their occupation at the time of the study? | Research Assistant (8) |
| Gender | Was the researcher male or female? | Female (N/R) |
| Experience and training | What experience or training did the researcher have? | Trained (8) |
| **Relationship with participants** | | |
| Relationship established | Was a relationship established prior to study commencement? | No (N/R) |
| Participant knowledge of the interviewer | What did the participants know about the researcher? e.g. personal goals, reasons for doing the research | Due to our purposive sampling method, and previous collaborations with participants in the field of preclinical patient engagement, participants had general personal and professional knowledge of the interviewer (9) |
| Interviewer characteristics | What characteristics were reported about the interviewer/facilitator? e.g. Bias, assumptions, reasons and interests in the research topic | Participants were informed that interviewers were interested in facilitating preclinical patient engagement (9) |
| **Domain 2: Study Design** | | |
| **Theoretical framework** | | |
| Methodological orientation and Theory | What methodological orientation was stated to underpin the study? e.g. grounded theory, discourse analysis, ethnography, phenomenology, content analysis | Thematic content analysis (8) |
| **Participant Selection** | | |
| Sampling | How were participants selected? e.g. purposive, convenience, consecutive, snowball | Purposive sampling (7) |
| Method of approach | How were participants approached? e.g. face-to-face, telephone, mail, email | Email recruitment (7) |
| Sample size | How many participants were in the study? | Table 1 |
| Non-participation | How many people refused to participate or dropped out? | One patient interview was excluded because it was determined they had not engaged in preclinical research (10) |
| **Setting** | | |
| Setting of data collection | Where was the data collected? e.g. home, clinic, workplace | Private room within the research institute or pre-admission unit (NR) |
| Presence of non-participants | Was anyone else present besides the participants and researchers? | Yes, caregivers (Table 1a) |
| Description of sample | What are the important characteristics of the sample? e.g. demographic data, date | Tables 1a-b (11) |
| **Data Collection** | | |
| Interview guide | Were questions, prompts, guides provided by the authors? | Interviewers used guides (7, Supplemental Files 4-6) |
|  | Was it pilot tested? | Yes (7,9) |
| Repeat interviews | Were repeat interviews carried out? If yes, how many? | No (N/A) |
| Audio/visual recording | Did the research use audio or visual recording to collect the data? | Yes (8) |
| Field notes | Were field notes made during and/or after the interview or focus group? | No (N/R) |
| Duration | What was the duration of the interviews or focus group? | Table 1a-b |
| Data saturation | Was data saturation discussed? | Yes (8) |
| Transcripts returned | Were transcripts returned to participants for comment and/or correction? | N/A |

| **Domain 3: Analysis and findings** | | |
| --- | --- | --- |
| **Data Analysis** | | |
| Number of data coders | How many data coders coded the data? | 19 (in duplicate)  -10 |
| Description of the coding tree | Did authors provide a description of the coding tree? | Yes (8, 10) |
| Derivation of themes | Were themes identified in advance or derived from the data? | From data (8) |
| Software | What software, if applicable, was used to manage the data? | NVivo11  -19 |
| Participant checking | Did participants provide feedback on the findings? | No (N/A) |
| **Reporting** | | |
| Quotations presented | Were participant quotations presented to illustrate the themes / findings? Was each quotation identified? e.g. participant number | No (N/A) |
| Data and findings consistent | Was there consistency between the data presented and the findings? | Yes (Table 2) |
| Clarity of major themes | Were major themes clearly presented in the findings? | Yes (Table 2) |
| Clarity of minor themes | Is there a description of diverse cases or discussion of minor themes? | Yes (Table 2) |
