## Supplementary Files - Identifying Challenges and Enablers to Engaging Patients in Preclinical Laboratory Research. for "Identifying Challenges and Enablers to Engaging Patients in Preclinical Laboratory Research": Additional File 3_GRIPP2.docx

**Additional File 3: Guidance for Reporting Involvement of Patients and the Public (GRIPP2) Short Form**

| **Section and Topic** | **Item** | Page number(s) |
| --- | --- | --- |
| 1: Aim | The aim of patient engagement was to ensure all aspects of the study included patient partner input. This was particularly important as our study team wanted to ensure the study design, conduct, and analysis aligned with the priorities and perspectives of researchers *and* patient partners (i.e., the target audience of our study results). | 9 |
| 2: Methods | The study team included four patient partners. Patient partner DPR led this group and co-facilitated regular study team meetings with lead researcher MML. Patient partners contributed to multiple study phases, including initial grant development (they were recognized as co-principal applicants [DPR] and co-applicants), interview guide development, protocol development for ethics applications, piloting the interview guide, data interpretation, and manuscript authorship. | 9 |
| 3: Study results | Consensus on the five key themes was reached over the course of two meetings that included all patient partners and our researchers. Patient partner feedback helped contextualize findings, particularly around communication, value-setting, and cultural shifts required in research. Their perspectives also helped frame actionable implications for future preclinical engagement initiatives. | 11 |
| 4: Discussion and conclusions | Patient partners shaped the interpretation of the study results as described above. Their insights emphasize the importance of accessibility, relationship building, and early-stage collaboration. These are reflected in the Discussion of our manuscript. Patient partner feedback on the writing of this manuscript was elicited and incorporated into the final version (all patient partners have been listed as co-authors). | 10 and 23 |
| 5: Reflections/critical perspective | Involving patient partners throughout the research process, from grant development to dissemination, improved clarity of study materials and strengthened interpretation. Co-developing the interview guide and iteratively discussing emerging findings helped ensure alignment with patient priorities. However, it required time and intentional planning to balance diverse perspectives on our team and accommodate schedules. Building and maintaining trust was critical, especially in a preclinical context unfamiliar to some patient partners. Compensation practices following CIHR guidelines supported equity but required administrative coordination. Overall, the process was valuable. | 25-26 |
