## Supplementary Files - Identifying Challenges and Enablers to Engaging Patients in Preclinical Laboratory Research. for "Identifying Challenges and Enablers to Engaging Patients in Preclinical Laboratory Research": Additional File 4_Interview Guide.docx

**Patient Partner Interview Guide**

I wanted to start off by thanking you for taking the time to speak with me today. As discussed, our research team is currently working on creating a list of suggestions for involving patients in preclinical research, also known as basic science or laboratory research (which means research that is done before it gets to people, such as genetics research or research with animals). We’re interested in speaking with you about your experience working with researchers on a preclinical research project. It’s important to note that there are no ‘right’ or ‘wrong’ answers to these questions; we’re simply interested in your perspective and suggestions, to help better inform our list of suggestions and make sure it is useful to other patients, the public and researchers.

Today’s interview should take about 60 minutes, but please take as much time as you need, and do not hesitate to let me know if you would like to take a break. As discussed, and if still okay with you, our interview will be voice-recorded to ensure that all key points are properly documented. The only people who will listen to the audio file will be members of the research team, and your responses will be ‘anonymized’ so that no one will know what your specific answers were. This just means that all identifying information (e.g. your name or the names of others) that you mention will be removed from the written record of the interview.

If you wish to end the interview before I have asked all of the questions or if you wish to withdraw from the study and remove your responses you are free to do so, just let me know. Any questions before we start?

**Background Questions**

My first few questions are about your background, so that we can describe our interviewees as a group, however please feel free to skip questions if you are not comfortable answering them:

1. In a sentence or two, can you tell me what patient/public engagement in research means to you?

2. If you are comfortable sharing, can you tell me a little about your everyday life? Are you a student, do you work, are you a caregiver or are you retired?

3. If comfortable sharing, can you please state your age or the year you were born?

4. If you are comfortable sharing, can you please state your biological sex (male, female, prefer not to say) and the gender you identify with (e.g. male, female, non-binary, prefer to self-describe, prefer not to say)?

5. If you are comfortable sharing, do you live with a condition or are you a caregiver?

6. How long have you been involved in research? How many projects have you been a part of?

7. Can you tell me how you first became interested or involved in research?

**Core Questions**

Great, thank you very much for sharing. As mentioned, today we’re hoping to learn more about how you were involved in the [*insert project name if known*] project and what your experience was like.

1. To start, I was wondering if you could tell me how you heard about the [*insert project name if known*] project (e.g. how did you become involved)?

a) *Prompt (as applicable):* Did you already know members of the research team/ committee? // Had you attended previous research conferences?
b) *Prompt:* Was there anything in particular that drew you to becoming involved in this specific project?

2. What was your (first) meeting with the research team like?

a) *Prompt:* Was the meeting in-person or virtual (online)? Who was at the meeting?
b) *Prompt:* Did the research team have processes in place to bring you onto the team (for example: for training, for setting goals/expectations, specifying roles, etc.)?

3. Can you tell me a little bit more about how you were involved [*in/throughout*] the project?

a) *Prompt:* Can you tell me more about the types of activities that you were involved in?
b) *Prompt:* Did you have a specific role?
c) *Prompt (as applicable):* From the paper describing this project, I understand you [attended/took part in a conference/ focus group/survey]. Did your engagement extend past this event?
d) *Prompt (as applicable):* How long were you engaged in the project for (e.g. number of events, months or years)?
e) *Prompt (as applicable):* Were there other patient partners/public partners/committee members involved? If so, how many? What effect did other members have on your involvement? What was your relationship with the other partners/members?

4. What was the environment or atmosphere of the team like (e.g. at meetings, events)?

a) *Prompt:* Did you feel comfortable/uncomfortable at meetings?
b) *Prompt:* Did you feel that you could speak freely and share your thoughts and perspectives easily?
c) *Prompt:* Did you feel included or uninvolved?

5. How did you communicate with the research team (e.g. in-person, over e-mail, by phone, all of these)?

a) *Prompt:* Did this method of communication work well for you? Why or why not?
b) Prompt: Did the research team communicate with you clearly (for example did they use clear language, or did they use any confusing terms or acronyms)?

6. Did the research team allow for flexibility in your involvement?

a) *Prompt:* Did the research team ask you your preferences for what you would like to work on?
b) *Prompt:* Was the research team flexible in scheduling meetings?
c) *Prompt:* Was the research team flexible in the amount of time for activities? Did they let you know that you could take a step back/ take breaks if needed?

7. Did the research team provide you with any forms of support (e.g. a point of contact, resources, training, transportation, compensation for your time)? Did you find it helpful?

a) *Prompt (if applicable)*: Did you have to ask the research team for this, or was it offered up front?

8. Did the research team acknowledge your input/ ideas/ contributions?

a) *Prompt (as applicable):* How did the research team acknowledge your input/ ideas/ contributions? For example, did the research team explain how they would include your ideas and feedback in the project or did they explain why they could not include your ideas and feedback? Did they thank you for your participation?
b) *Prompt (as applicable):* How did you feel about the way your contributions were acknowledged? Do you think your contributions could have been acknowledged in a different way?

9. Did the research team make an effort to provide you with updates or the study results?

a) *Prompt (as applicable):* How often did you connect with the research team? Did you receive updates even when you were not being asked to contribute to the project?
b) *Prompt (as applicable):* Did the research team try to maintain the partnership beyond the research project (e.g. invite you to participate in events or other projects, continue sending updates)?

10. Looking back, how was your overall experience (e.g. positive or negative experiences, both)?

a) Was there anything that helped contribute to positive experiences?
b) Was there anything that contributed to negative or challenging experiences?
a) If you did have a negative experience, how did you overcome it, or what would have helped to overcome it? Do you think these challenges can be overcome in future engagement activities/programs?
b) Were these challenges expected or unexpected?

11. Was engaging in this research project meaningful/worthwhile to you personally? Why or why not?

12. If you could go back, is there anything you would have done differently?

13. Is there anything the research team could have done differently?

14. Is there anything that might help to improve future experiences (for you or others)?

15. Were there any other activities or areas that you would have liked to have been involved in?

a) *Prompt:* For example, establishing partnerships, forming the research question, designing the project, seeking funding, submitting an application to the ethics board, during the experiment/ data collection, analysis of data, drawing conclusions, publishing the results, sharing the results with others and implementing the findings (e.g. knowledge translation), etc.

16. Have you, or do you plan to, partner with researchers again? Why or why not?

17. Is there anything else you would like to add?

Thank you very much for participating!

**Researcher Interview Guide**

I wanted to start off by thanking you for taking the time to speak with me today. As discussed, our research team is currently working on a framework of promising practices for engaging patients in preclinical research. We’re interested in speaking with you about your experience engaging patients in preclinical research. It’s important to note that there are no ‘right’ or ‘wrong’ answers to these questions; we’re simply interested in your perspective and suggestions, to help better inform our framework and make sure it is useful to other researchers, patients and the public.

Today’s interview should take approximately 60 minutes and will be audio-recorded just to ensure all key points are accurately documented. Your responses will be pseudonymized. As soon as we finish, I’ll assign your audio file a unique ID code and store it on the hospital server. The only people who will listen to the audio file will be authorized study personnel and the contracted transcriptionist. When the audio file is converted to text format we will ensure that any and all identifying information (for example, your name or names of other individuals that you use in the course of our discussion) is removed from the interview transcripts.

If you wish to end the interview before I have asked all of the questions or if you wish to withdraw from the study you are free to do so. Any questions before we start?

**Background/Demographic Questions**

1. What disease or areas of research do you work in (e.g. cancer, autism, rheumatic/musculoskeletal conditions, sepsis, neuromuscular disorders, Alzheimer’s disease/dementia, psychiatric genomics, rare diseases, other area, etc.)?

2. In a sentence or two, what does patient or public engagement in research mean to you?

3. Which option best describes you:

a) Early career researcher (have held a full time, independent research appointment, for a period of 0 to 5 years [60 months])
b) Mid-career researcher (assumed your independent research position 5-15 years ago)
c) Senior researcher (assumed your first independent research position more than 15 years ago)

4. If comfortable sharing, can you please state your age or the year you were born?

5. If you are comfortable sharing, can you please state your biological sex (male, female, prefer not to say) and the gender you identify with (e.g. male, female, non-binary, prefer to self-describe, prefer not to say)?

6. If comfortable, could you please share what influenced your decision to engage patients/ the public in your research?

**Core Questions**

Great, thank you very much for sharing. As mentioned, today we’re hoping to learn more about your experience engaging patients/ the public in your project [*insert project name if known*].

1. To start, I was wondering if you could tell me how you initiated engagement or recruited patient partners/the public?

a) *Prompt (as applicable):* Had you engaged with this patient organization/ committee/ patient/ community member before?

2. What was your (first) meeting with the partners like?

a) Were there processes in place (e.g. for onboarding, for setting goals/expectations, specifying roles, etc.)?
b) *Prompt (if applicable)*: How did you develop these processes? Were there any resources that helped you develop or conduct these processes?

3. Can you tell me a little bit more about how patients/the public were engaged?

a) *Prompt:* What types of activities were patient/public partners involved in?
b) *Prompt:* Did patients/the public have a specific role?
c) *Prompt:* Were these activities/ roles pre-determined? Did they evolve over the course of the project?
d) *Prompt (as applicable):* From the paper describing this project I understand partners/the public [attended/took part in a conference/ focus group/ survey]. Did their engagement extend past this event?
e) *Prompt (as applicable):* How long did patient/public involvement last (e.g. number of events, months or years)?
f) *Prompt:* How many patient/public/committee members were involved?
g) *Prompt:* Did the team receive funding specifically for the engagement component of the project (e.g. was patient engagement included in the grant budget)? If so, can you share which organization(s) provided funding?
h) *Prompt:* Were patient/public partners compensated or did they receive an honorarium (e.g. gift card) or reimbursement (e.g. travel, accommodation)?

4. What was the environment or atmosphere of the team like during meetings/ events?

a) *Prompt:* Was there a chance for patient/ public partners to be introduced to all team members?
b) *Prompt:* Were you able to make sure everyone had a chance to share thoughts and ideas throughout meetings?

5. How did you communicate with patient partners/the public (e.g. in-person, over e-mail, on the phone or all of the above)?

a) *Prompt:* Did this method of communication work well for you? Why or why not?
b) *Prompt:* How did you find explaining your research to patient partners/the public? Easy or difficult?

6. Looking back, how was your overall experience (e.g. positive or negative experiences, both)?

a) Was there anything that helped contribute to positive experiences?
b) Was there anything that contributed to negative or challenging experiences (e.g. barriers)?
a) If you did experience a challenge, how did you overcome it, or what would have helped to overcome it? Do you think this challenge can be addressed in future patient engagement activities/ programs?
b) Were these challenges expected or unexpected?

7. Did you find engaging patients/the public in your research project meaningful/worthwhile to you personally? Did it help with your research objectives and overall project? Why or why not?

8. If you could go back, is there anything you would have liked to have done differently? Why and how?

9. Are there any important structure or process considerations to take into account when engaging patients/the public (e.g. for onboarding, for setting goals/expectations, specifying roles)?

10. Are there any important institutional or organizational considerations to take into account when engaging patients/ the public (e.g. hospital approval, ethics board approval, patient organization approval)?

11. Is there anything that might help you or others to engage patients/the public in your future preclinical work?

a) As an example, how do you think social media could help you to engage patients/ the public in your research?
b) Did you come across any helpful guidance, training or funding opportunities that might be helpful to others?

12. To help us better inform our framework, we were wondering if you have any other ideas as to how patients could be engaged throughout the various stages of preclinical research?

a) *Prompt:* Identifying and prioritizing research areas
b) *Prompt:* Hypothesis generation
c) *Prompt:* Experimental design
d) *Prompt:* Conducting experiments
e) *Prompt:* Results synthesis & data analysis
f) *Prompt:* Interpretation
g) *Prompt:* Dissemination
h) *Prompt:* Validation
i) *Prompt:* Re-calibration of research question

13. Have you, or do you plan to, engage patients/ the public again in your future work? Why or why not?

14. Anything else you would like to add?

Thank you for participating!
