## Supplementary Files - Identifying Challenges and Enablers to Engaging Patients in Preclinical Laboratory Research. for "Identifying Challenges and Enablers to Engaging Patients in Preclinical Laboratory Research": Additional File 5_PEBS Codebook.docx

**PEBS Interview Study Codebook (*Developed from Partner Interviews #1-3, Researcher #1-2)**

| **Code** | **Sub-code** | | **Definition** |
| --- | --- | --- | --- |
| **REASONS FOR ENGAGEMENT** | | | |
| Purpose of Engagement |  | | General view or description of why patients should be engaged in research, or when they respond to the question ‘what does patient engagement in research mean to you?’. May also include when a participant describes why patient engagement is important/ why others should. |
| Objective | General | | Planned aim of the patient engagement initiative (e.g. getting researchers to understand patients, sharing perspectives) |
|  | Better understanding and communication of research | | Program focuses on improving the communication skills of researchers, or to inform and teach patients about research (e.g. sharing what research is ongoing/ explaining research concepts to patients/ awareness of opportunities). |
|  | Change scientific culture | | The program aims to change the scientific community’s view of ***patient engagement*** in research (e.g. want to train the next generation of researchers). |
|  | Build relationships | | To make connections between patients and researchers, so that there is a strong relationship (e.g. for future engagement). Can include building a patient partner community. |
|  | Develop guidance on engagement | | The objective of the program is to develop guidance, frameworks, and/or research patient engagement in basic science. For example, the team was also interested in identifying barriers and facilitators to patient engagement. |
| [Partner/ Researcher] Motivation  (*Select appropriate option when coding) | General | | Motivation for engagement; why the partner engages in research. |
|  | Personal interest | | Personal interest in condition (e.g. link to disease, bad healthcare experience), in study/project, in patient engagement, or being involved in research in general (e.g. intellectually stimulating). |
|  | Altruism | | E.g. To help improve research for other patients; ethically the right thing to do. |
|  | Change scientific culture | | The participant expresses the desire to change the scientific community’s view of patient engagement in research (e.g. want to train the next generation of researchers). |
| **ROLES** | | | |
| [Partner/ Researcher] Role  (*Select appropriate option when coding) | General | | How the participant contributes to the engagement program. Typically described in one to two words. |
|  | Patient voice | | Role to provide patient perspective in research and to represent the patient voice or point of view. May include advisory/committee members that advise on research, grants, faculty hiring (e.g. interview panel, steering committee). |
|  | Leader | | In charge of key duties in engagement program. |
|  | Facilitator | | Helps facilitate communication or partnership between patients and researchers, helps to onboard, mentor and train patient partners, or helps to facilitate activities. Can also be a point of contact for questions or concerns. Synonyms: Mentor, liaison, point of contact. |
|  | Champion | | Champions patient engagement in research/ engagement initiative. May also be described as encouraging. |
| **LOGISTICS** | | | |
| Logistics | General | | Describes team organization in terms of team members, resources and infrastructure. |
|  | Number of partners | | Mention of the number of patient partners involved in the project or program. Can also use this to flag number of patients/caregivers in audience at an educational/awareness seminar. |
|  | Timeline | | Description of length of project. |
|  | Mode of communication | | How the researchers and partners communicate. Ex. Email, phone, face to face meetings. Also include mention of meeting logistics, such as the frequency of meetings, scheduling meetings, where and when meetings took place. Also include participant preferences for mode of communication and correspondence here. |
|  | Program funding | | Describe funding for the specific program. For example, how or where they applied for funding, or whether or not they received funding. |
|  | Compensation style and support | | How the partners were compensated for their involvement. E.g. Voluntary, reimbursement, paid/contract. Also include non-financial supports. E.g. Point of contact, transportation, mentors/facilitators. |
|  | Partner population | | Use this code to flag the unique population being engaged, e.g. pediatric, geriatric, public, experienced chemotherapy, etc. |
|  | Research team | | Description of individual members on the research team or involved in the project |
| **PROCESSES** | | | |
| Process | General | | Process for patient partner engagement. |
|  | Initiation | | Describe how partnership started (e.g. who reached out, who they reached out to). How they met or connected. |
|  | Training and onboarding | | Description of how training or onboarding for patient partners and/or researchers was developed or conducted. May also include the process for defining the proper terminology and clear language (scientific, engagement, etc.) and how partners can indicate when researchers are using unclear language. |
|  | Development of roles, responsibilities, activities and materials | | Describe how team member roles, responsibilities, activities and materials are developed. E.g. Roles evolved over time or were co-developed; participant described that the partners helped to pilot materials/presentations; describe setting expectations or goals. |
|  | Team building | | Describe processes or activities completed to build relationships between partners and researchers. Or participant describes the importance of building a relationship with the partner(s) (e.g. getting to know them). |
|  | Evaluation and feedback | | Indicates that the team asks partners /researchers for feedback on *the engagement program* or conducts an evaluation of the engagement program. Or describes how this is done. |
|  | Dissemination | | Describe how the team shares research or program approach with others. |
| **STRUCTURES/ ENVIRONMENT** | | | |
| Team Structure | General | |  |
|  | Leadership | | Participant describes the team structure or patient-researcher relationship (e.g. if there was a hierarchical structure with leaders, researcher-partner co-leadership, if there were facilitators, if partners were broken up into groups, etc.). |
|  | Connection or collaboration | | Use this code to flag when participant describes activities associated with involvement in a patient organization. This may be just a connection with the patient partner or a researcher-charity/organization collaboration. |
|  | Pre-existing relationships | | The researchers and patient partners previously knew each other from a previous engagement activity. Or used their network to identify partners. |
| Structures | General | | Describes a characteristic, feature, or element of the engagement program (e.g. how the program is set-up). |
|  | Flexible and fluid | | Describe how engagement is flexible (e.g. allow partners to join in on activities as they please, partners brought in as needed). |
|  | Engage early-career | | Program engages young researchers (e.g. students, trainees, early career scientists). |
|  | Engage patients early | | Researchers engage patients early in the research process (e.g. conception of project). |
|  | Ongoing program | | The engagement initiative is structured as a ‘program’ involving multiple events/objectives/networks. Versus a ‘non-ongoing program’ (e.g. one time study). |
|  | Interactive activity or material | | Explicit mention that they tried to develop an interactive activity to engage/entertain partners. E.g. Interactive teaching material, learning workshop |
| Environment or Team Dynamic | General | | Description of the team atmosphere during the collaboration. May be explicit (e.g. the team was friendly, easy-going, etc.) or implicit (e.g, We had a lot of fun working together) |
|  | Open and inclusive | | Participant describes team ***atmosphere*** as open, welcoming, inclusive, comfortable, respectful, etc. May also discuss considerations for making the environment open and inclusive (i.e. including siblings of patients). |
|  | Informal/formal | | Describes the team environment as either formal (e.g. serious) or informal (fun, more laid back). |
| Communication | General | |  |
|  | Open line of communication | | Participant describes example of how the team developed or encouraged learning and an open line of communication, or the importance of this. For example, the participant may describe defining activities, roles and expectations to partners (e.g. what activities are involved, time commitment, compensation). |
|  | Lack of communication | | Participant describes an example of when there were communication problems. |
| **RESPONSIBILITIES & ACTIVITIES** | | | |
| Partner Responsibilities and  Activities | General | | Describes actions/activities that partner undertook over the course of the project (e.g. disseminating articles to others). May also include other activities not explicitly described as engagement, such as lobbying and advocating. |
|  | Research standards, design and methodology | | Partners are expected to adhere to appropriate/ rigorous research standards or methods. Or participant comments that patients should not provide input on scientific methods. |
|  | Developing documents | | The partner’s activities involve creating or reviewing documents to provide the patient perspective and ensure language is appropriate. For example: research proposals, grant applications, manuscripts, summaries, abstracts or any other documents. |
|  | Training and support | | E.g. provide training or support to other patient partners or to researchers. May also be described as interpreting, translating or acting as a bridge between partners and researchers. |
|  | Involved in recruitment | | Planning/helping recruit patients. Can involve the direct recruitment of other patients, being involved with the recruitment strategy or providing feedback on recruitment initiatives. |
|  | Coordinate and plan meetings or programs | | Partner helps to develop, plan or coordinate the engagement program or meetings (e.g. logistics, activities). |
|  | Providing feedback | | Provide feedback on the patient partner program, research, or identifying research priorities. For example, the partner’s activities involve listening and providing feedback on researcher presentations to ensure communication is clear (e.g. communication). Or to ensure researcher’s ideas and rationale are clear (e.g. ask questions). |
|  | Presenting | |  |
| Researcher Responsibilities and Activities | General | | Example: Sharing about their research, raising awareness. |
|  | Training and support | | E.g. provide training to patient partners or to other researchers. |
|  | Present scientific concepts ***accessibly*** | | Present research to patients or public and ensure the material can be easily understood. Can also be described as educating or teaching patients/public. |
|  | Facilitate discussion | |  |
| **TRAITS, EXPERIENCES AND VIEWS** | | | |
| Experience | General | | Can include other advocacy activities. |
|  | Past experience | | Participant describes a past engagement experience or notes that they have engaged patients/ engaged in research before. E.g. Partner was involved in a research review panel. Previously labelled as ‘Level of experience (with engagement/research)’. |
|  | Positive experience | | Participant describes a positive patient engagement experience. E.g. patient partner feels valued, plan to engage with patients/researchers again, etc. |
|  | Negative experience | | Participant describes a negative patient engagement experience. E.g. patient partner does not feel valued. |
|  | Newness of patient engagement | |  |
| Partner Traits | General | | Include traits and self-concept codes. |
|  | Enthusiastic and motivated | | E.g. Partner indicates they wished they had started earlier in their career, expresses commitment to engagement/program. |
|  | Established | | May also be described as respected within the community. |
|  | Skills and expertise | | When a participant describes a skill or expertise. For example, that they work in a medical profession, that they are used to public speaking, or that they are familiar/experienced with academics/students. |
|  | Assertive and confident | | Participant expresses willingness to speak up or make their voice heard. Note: May be viewed as either a positive or negative trait (e.g. confident or undiplomatic). |
| Researcher Trait | General | |  |
|  | Collaborative | | Participant makes reference to the researcher(s) being a good facilitator, partner or collaborator, or that this is a helpful characteristic. |
|  | Less social | | May also be described as ‘Not comfortable or familiar speaking to patients/public’. |
|  | Enthusiastic and motivated | |  |
|  | Established | | May also be described as respected within the community. |
|  | Siloed within research area | |  |
| Partner Preferences | General | | Preferences or expectations. This code will typically be linked to another code to specify the category (e.g. Process: Training, Structure: Engage partners early, Cultural/Country/Institution: Funding Scheme). |
|  | Engagement features | | Participant describes preferences for engagement. For example: Partners preferred being involved in one project as opposed to multiple, direct engagement with the PI/researcher, interactive activities, being engaged early, or deciding on the length and intensity of involvement. |
|  | Personal interest | | Prefer to work on a project focusing on a condition for which they have a personal link (e.g. have experienced this condition/ therapy, are a caregiver for this condition), or express that they need to believe in the project. |
|  | Compensation, acknowledgement and support | | Participant expresses their preference, view or expectation for compensation, acknowledgement or support. For example, they express that they do not expect compensation. Or describe their preferences for the accompanying activities (e.g. travel, local vs nation programs, administrative burden, etc.). |
| Researcher Preferences | General | | Preferences or expectations. This code will typically be linked to another code to specify the category (e.g. Process: Training, Structure: Engage partners early, Cultural/Country/Institution: Funding Scheme). |
|  | Engagement features | | Participant describes preferences for engagement. For example: Deciding on the length of engagement. |
|  | Measure impact | | Note that they are interested in or would like to measure the impact of engagement (e.g. overall, on the research, etc.). |
| Perceptions and Views | General | | Views about research or patient engagement in general |
|  | Perception of partner | Not comfortable with scientists | For example, participant mentions that qualifications can be intimidating. |
|  |  | Not interested in preclinical research |  |
|  |  | Are interested and intelligent |  |
|  | Perception of researchers | Not comfortable or interested in working with patients | May also include describing researchers as isolated within the lab or isolated from real patients. May also be described as ‘stuck in their ways’. |
|  |  | Accepting of patient partners | Partner makes reference to obtaining researcher or clinician acceptance of their role in research. |
|  |  | Provision of medical advice | Participant makes note that patients sometimes want to ask medical questions, but that this is not appropriate for preclinical researchers/students. |
|  | Views on industry | | Participant makes reference to ‘industry’ (e.g. practices, engagement). |
|  | Ethical considerations and views | | Participant highlights ethical considerations or concerns around engaging patients in research, or animal use. |
| **CHALLENGES & CONSIDERATIONS** | | | |
| Challenge | General | | Examples: Limitations in research. |
|  | Finding interested researchers or partners | | Participant describes that they had issues in finding or reaching out to researchers or partners. They may also describe as there being no clear ‘entry point’ for interested patient partners. |
|  | Explaining or understanding scientific concepts | | Participant expresses that it is either difficult for researchers to explain science to patients, or for patients to understand complex scientific concepts. For example, the participant may express that patients typically do not have knowledge of basic scientific concepts. |
|  | Diversity | | Participant mentions difficulties in ensuring engagement was equitable for all participants. For example, participant mentions the team was not able to involve small children or patients with complex needs. |
|  | Maintaining or sustaining the relationship | | Participant describes difficulties in maintaining the collaboration or relationship between partners/researchers at the end of the project. |
| Considerations | General | |  |
|  | Pediatric population | | Important things to keep in mind when engaging with a pediatric population (e.g. whether their parents or siblings should be present). |
|  | Partner population | | Noting the importance of tailoring engagement to the partner group or other cultural considerations when engaging with these partners. |
|  | Location or environment matters | | Participant notes that it is important to consider the location (or that they did this for their program). E.g. Neutral to reduce intimidation, easily accessible, etc. May also note considerations if team members are geographically dispersed. |
|  | Patient burden and availability | | Patient symptoms or side effects from condition or competing priorities. Also includes burden on patient, such as time commitments, or that the team took this into consideration (e.g. made it easier for the patient partner). |
|  | Researcher time, workload, and competing priorities | | Mention time constraints, deadlines, or other priorities (e.g. family, work). |
|  | Patient partner dropouts | | Mention that patients sometimes need to take a break or step back, or that over time it can be expected that patient partners may drop out. |
|  | Social media pros and cons | | May include pros and cons of social media to engage with patients or the public. |
|  | Diversity | | Participant notes that it’s important to have a diverse group of patients, partners, or researchers. |
|  | Confidentiality or COI | | Partner mentions concerns around sharing confidential information, or conflicts of interests between team members, patients, or institutions regarding patient engagement or their project. |
| Cultural/Country/Institutional Factors | General | | Cultural, country and institutional factors for patient engagement. E.g. institutional practice or mandate (including around communication with the public), country norms in engagement |
|  | Funding scheme | | When generally speaking about the system for funding. For example, that there are few/many funding opportunities. |
|  | Structures for encouraging | | E.g. University course, interview panel for faculty includes patient, grant requires patient engagement, the Research Ethics Board provides an ethics statement on patient engagement. |
|  | Traditional culture of the scientific community | | E.g. Isolated from patients, focus on ‘media worthy science’, publish or perish culture, no direct career benefits (or that they would be helpful) |
|  | Lack of guidance | | Participant suggests there is a lack of guidance on engaging patients in preclinical research, or that guidance would be helpful. |
|  | Understanding the value of patient engagement | | General understanding or acceptance (or **not**) within the scientific community. For example: Institutional culture promotes the value of PE, perception that PE not valued academically, superficial engagement for public relations, etc. |
|  | Entry point for engagement | | The institution provides an access point for engagement (e.g. hospital or organization provides access to partners). |
| **OUTCOMES** | | | |
| Benefits and Outcomes  *Specify if benefit to researcher or partner if clear | General | | Participant makes note of an outcome or benefit from the patient engagement. Should specify whether it is a benefit to the partner, researcher or to the program. More general than ‘Motivation’ codes (why they engage). |
|  | Expanding to other opportunities | | Patient engagement program led to new opportunities, such as work experience for students, additional talks, or built relationships with individuals due to the engagement. |
|  | Learning and skill development | | When patients or researchers mention that the engagement led to learning. This could mean they learned something new, obtained a better understanding of already learned concepts, or gained a different/new perspective. May also include mention that they developed new or existing skills (e.g. Communication skills, public speaking), gained additional expertise, or intra-personal improvements (e.g. More confident). |
|  | Meaningful and rewarding | | When patients or researchers feel the engagement was personally meaningful, rewarding, worthwhile. They may discuss feeling valued and acknowledged. |
|  | Procedural or research | | Outcomes of the engagement that benefited the research endeavors. This could include helping recruit patients, being awarded a research grant, helping disseminate research, etc. |
| **TERMINOLOGY/ LANGUAGE USED BY PARTICIPANT** | | | |
| Terminology |  | | Use the ‘Terminology’ code to flag terms associated with patient engagement. Will be helpful in highlighting differences across teams. For example: Patient Research Partners (PRPs), participatory research, tokenism, patient advocate, involvement, outreach, community member. |
| **OTHER** | | | |
| Demographics |  | | Age, gender, everyday life, etc. |
| Overlap with clinical experience |  | | Flag when participant describes example pertaining to engagement in clinical research project (e.g. clinical trial). |
| Existing or other engagement resources |  | | Use this code to flag when a participant describes use, development or existence of other patient engagement resources (which may be helpful in the development of the framework). May also include resources used to train or onboard partners or researchers, for example: online content for basic science knowledge. May also include guidance from other fields (e.g. product design, social science, etc.). |
